# Estimating the COVID-19 epidemic trajectory and hospital capacity requirements in South West England: a mathematical modelling framework

**DOI:** 10.1101/2020.06.10.20084715

**Authors:** Ross D. Booton, Louis MacGregor, Lucy Vass, Katharine J. Looker, Catherine Hyams, Philip D. Bright, Irasha Harding, Rajeka Lazarus, Fergus Hamilton, Daniel Lawson, Leon Danon, Adrian Pratt, Richard Wood, Ellen Brooks-Pollock, Katherine M.E. Turner

## Abstract

**Objectives:** To develop a regional model of COVID-19 dynamics, for use in estimating the number of infections, deaths and required acute and intensive care (IC) beds using the South West of England (SW) as an example case.

**Design:** Open-source age-structured variant of a susceptible-exposed-infectious-recovered (SEIR) deterministic compartmental mathematical model. Latin hypercube sampling and maximum likelihood estimation were used to calibrate to cumulative cases and cumulative deaths.

**Setting:** SW at a time considered early in the pandemic, where National Health Service (NHS) authorities required evidence to guide localised planning and support decision-making.

**Participants:** Publicly-available data on COVID-19 patients.

**Primary and secondary outcome measures:** The expected numbers of infected cases, deaths due to COVID-19 infection, patient occupancy of acute and IC beds and the reproduction (“R”) number over time.

**Results:** SW model projections indicate that, as of the 11^th^ May 2020 (when ‘lockdown’ measures were eased), 5,793 (95% credible interval, CrI, 2,003 – 12,051) individuals were still infectious (0.10% of the total SW England population, 95%CrI 0.04 – 0.22%), and a total of 189,048 (95%CrI 141,580 – 277,955) had been infected with the virus (either asymptomatically or symptomatically), but recovered, which is 3.4% (95%CrI 2.5 – 5.0%) of the SW population. The total number of patients in acute and IC beds in the SW on the 11^th^ May 2020 was predicted to be 701 (95%CrI 169 – 1,543) and 110 (95%CrI 8 – 464) respectively. The R value in SW England was predicted to be 2.6 (95%CrI 2.0 – 3.2) prior to any interventions, with social distancing reducing this to 2.3 (95%CrI 1.8 – 2.9) and lockdown/ school closures further reducing the R value to 0.6 (95CrI% 0.5 – 0.7).

**Conclusions:** The developed model has proved a valuable asset for local and regional healthcare services. The model will be used further in the SW as the pandemic evolves, and – as open source software – is portable to healthcare systems in other geographies.

**Future work/ applications:**

i. Open-source modelling tool available for wider use and re-use.
ii. Customisable to a number of granularities such as at the local, regional and national level.
iii. Supports a more holistic understanding of intervention efficacy through estimating unobservable quantities, e.g. asymptomatic population.
iv. While not presented here, future use of the model could evaluate the effect of various interventions on transmission of COVID-19.
v. Further developments could consider the impact of bedded capacity in terms of resulting excess deaths.

## Introduction

Since the initial outbreak in 2019 in Hubei Province, China, COVID-19, the disease caused by Severe Acute Respiratory Syndrome Coronavirus-2 (SARS-CoV-2), has gone on to cause a pandemic [1]. As of 11^th^ May 2020, the Centre for Systems Science and Engineering at Johns Hopkins University reports over 4,000,000 confirmed cases and 250,000 deaths globally [2]. National responses to the outbreak have varied; from severe restrictions on human mobility alongside widespread testing and contact tracing in China [3] to the comparatively relaxed response in Sweden, where lockdown measures have not been enacted [4]. In the UK, advice to socially distance if displaying symptoms was given on the 15^th^ March, while school closures and ‘lockdown’ measures were implemented from 23^rd^ March onwards [5].

Mathematical modelling has been used to predict the course of the COVID-19 pandemic and to evaluate the effectiveness of proposed and enacted interventions [6–11]. These models have been predominantly aimed at the national level and have largely been based on epidemiological and biological data sourced from the initial epidemic in Wuhan, China [12] and the first large outbreak in Lombardy, Italy [13].

In the UK, the epidemic escalated most rapidly in London [14] and the majority of national modelling is seemingly driven by the trends in London, due to its large case numbers and large population. One of the key issues facing NHS authorities is planning for more localised capacity needs and estimating the timings of surges in demand at a regional or healthcare system level. This is especially challenging given the rapidly evolving epidemiological and biological data; the changes in COVID-19 testing availability (e.g. previously limited and changing eligibility requirements); the uncertainty in the effectiveness of interventions in different contexts; significant and uncertain time-lags between initial infection and hospitalisation or death; and different regions being at different points in the epidemic curve [9]. South West England (SW) is the region with the lowest number of total cases in England (as of 11^th^ May 2020), lagging behind the national data driven by the earlier epidemic in London [9, 14].

COVID-19 results in a significant requirement for hospitalisation, and high mortality amongst patients requiring admission to critical care (particularly amongst those requiring ventilation) [15, 16]. In the SW, the population is on average older than in London [17] and is older than the UK as a whole (Table S1). Older age puts individuals at elevated risk of requiring hospital care [18–20]. Consequently, we might expect higher mortality and greater demand for beds in the SW than estimations output from national models that may lack such granularity or risk sensitivity.

However, the SW’s first case occurred around 2 weeks later than the first UK case [14]; perhaps implying that the local SW epidemic may be more effectively controlled due to a lower number of baseline cases (than the national average) at the time national interventions were implemented, as well as reduced transmission due to rurality. This sub-national analysis can support in mapping the local epidemic, planning local hospital capacity outside of the main urban centres, and ensuring effective mobilisation of additional support and resources if required. Should demand be lower than expected, reliable forecasts could facilitate more effective use of available resources through re-introducing elective treatments (that had initially been postponed) and responding to other, non-COVID-19 sources of emergency demand.

In this study, taking into account the timeline of UK-wide non-pharmaceutical interventions (social distancing, school closures/lockdown) we illustrate use of our model in projecting estimates for the expected distributions of cases, deaths, asymptomatic and symptomatic infections, and demand for acute and intensive care (IC) beds. We present the model trajectories for SW England using publicly-available data.

## Methods

We developed a deterministic, ordinary differential equation model of the transmission dynamics of COVID-19, including age-structured contact patterns, age specific disease progression and demand for hospitalisation, both to acute and IC. We then parameterised the model using available literature and calibrated the model to data from the SW. The model is readily adapted to fit data at sub-regional (e.g. Clinical Commissioning Group), regional or national level. Key assumptions of the model are summarised in the supplementary information.

The model was developed in R and all code and links to source data are freely available (*github.com/rdbooton/bricovmod*). The model is coded using package *deSolve*, with contact matrices from package *socialmixr*, and sampling from package *lhs*.

### Model structure

The stages of COVID-19 included within this model are ***S***-susceptible, ***E***-exposed (not currently infectious, but have been exposed to the virus), ***A****-* asymptomatic infection (will never develop symptoms), ***I***-symptomatic infection (consisting of pre-symptomatic or mild to moderate symptoms), ***H***-severe symptoms requiring hospitalisation but not IC, ***C***-very severe symptoms requiring IC, ***R***-recovered and ***D****-* death. The total population is ***N = S + E + A + I + H + C + R + D*** (Figure 1).

**Figure 1:**
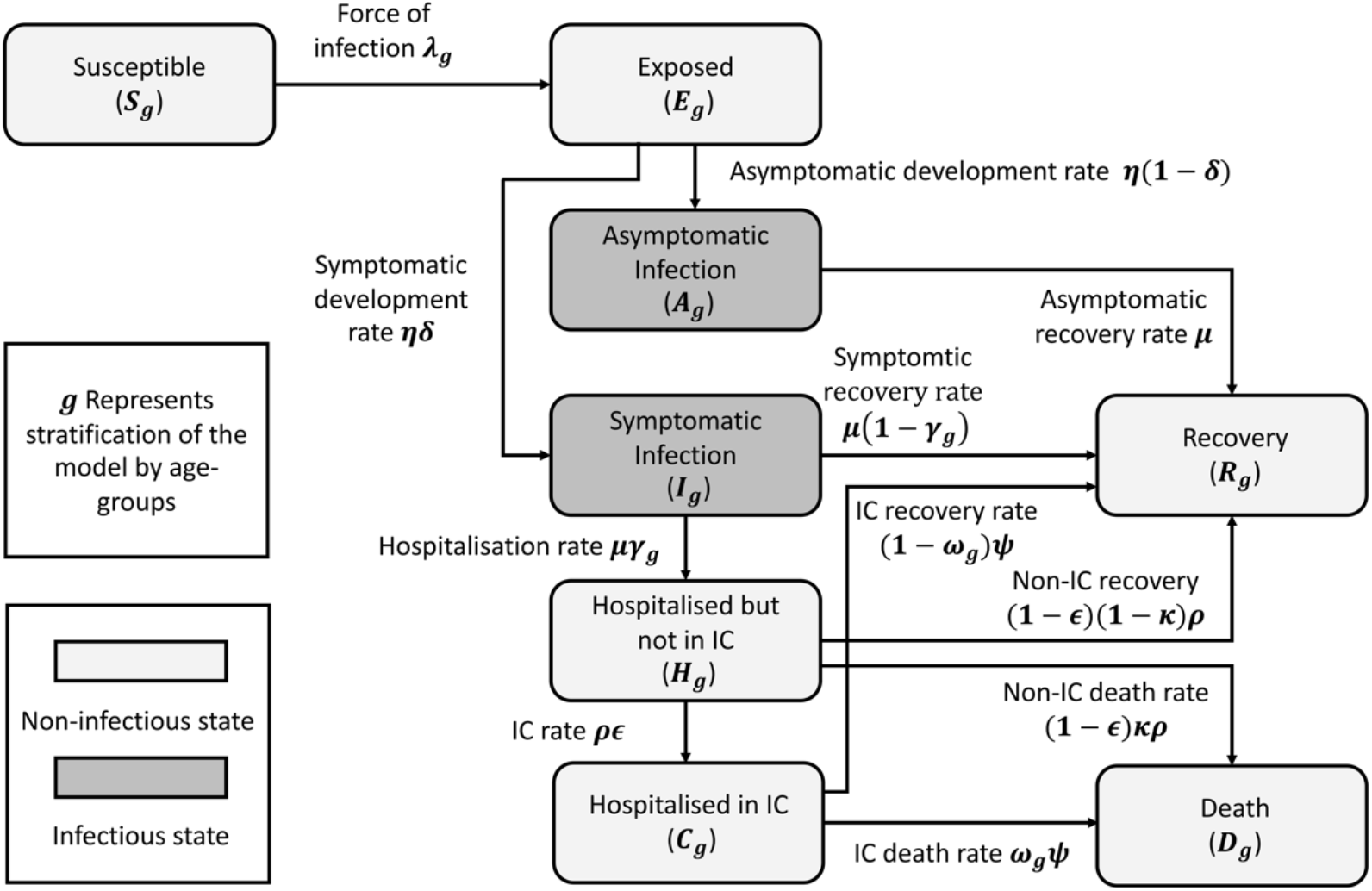
Compartmental flow model diagram depicting stages of disease and transitions between states. Asymptomatic infection represents the number of people never showing symptoms, while symptomatic infection includes all those who show pre-symptomatic / mild symptoms to those who show more severe symptoms (pre-hospitalisation). Those who are hospitalised first occupy a non-IC bed (acute bed) after which they can either move into IC, recover or die. Those in IC can either recover or die at an increased rate compared to those in acute beds. This model does not capture those deaths which occur outside of hospital as a result of COVID-19.

Each compartment ***X_g_*** is stratified by age-group (0-4, 5-17, 18-29, 30-39, 40-49, 50-59, 60-69, ≥70) where ***X*** denotes the stage of COVID-19 (S,E,I,A,H,C,R,D) and ***g*** denotes the age group class of individuals. Age groups were chosen to capture key social contact patterns (primary, secondary and tertiary education and employment) and variability in hospitalisation rates and outcomes from COVID-19 especially in older age groups. The total in each age group is informed by recent Office for National Statistics (ONS) estimates [21].

Susceptible individuals become exposed to the virus at a rate governed by the force of infection ***λ_g_***, and individuals are non-infectious in the exposed category. A proportion ***δ*** move from exposed to symptomatic infection and the remaining to asymptomatic infection, both at the latent rate ***η***. Individuals leave both the asymptomatic and symptomatic compartments at rate ***μ***. All asymptomatic individuals eventually recover and there are no further stages of disease: the rate of leaving the asymptomatic compartment is therefore equivalent to the infectious period, ***μ***. A proportion of symptomatic individuals ***γ_g_*** go on to develop severe symptoms which require hospitalisation, but not intensive care. Once requiring hospitalisation, we assume individuals are no longer infectious to the general population due to self-isolation guidelines restricting further mixing with anyone aside from household members (if unable to be admitted to hospital) or frontline NHS staff (if admitted to hospital). Individuals move out of the acute hospitalised compartment at rate ***ρ***, either through death, being moved to intensive care at rate ***∊***, or through recovery (all remaining individuals). A proportion ***ω_g_*** of patients requiring IC will die at rate ***ψ***, while the rest will recover.

The model (schematic in Figure 1) is therefore described by the following differential equations:

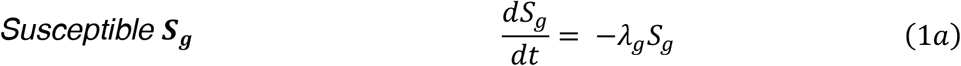

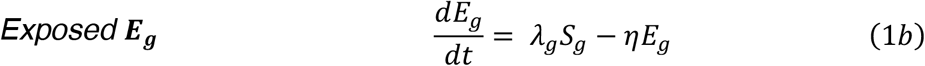

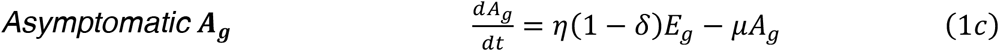

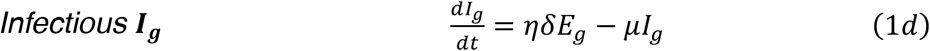

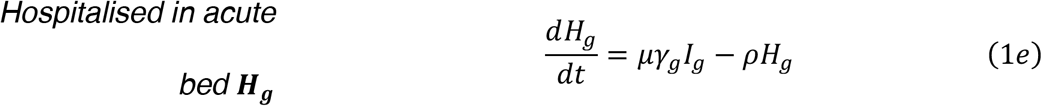

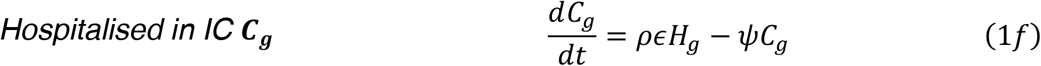

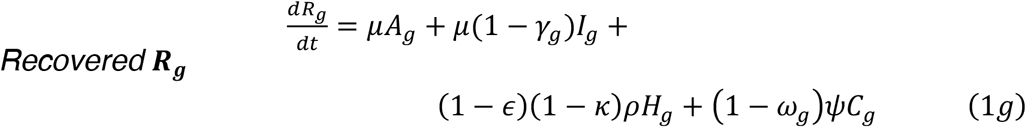

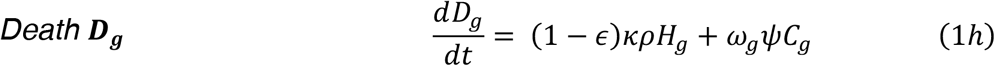

### Contact patterns under national interventions

We assume the population is stratified into pre-defined age groups with age-specific mixing pattern represented by a contact matrix ***M*** with an element of ***m_ij_*** representing the contacts between someone of age group ***i*** ∊ ***G*** with someone in age group ***j*** ∊ ***G***. The baseline contact matrix (with no interventions in place) is taken from the POLYMOD survey conducted in the United Kingdom [22]. The contact pattern may also be influenced by a range of interventions (social distancing was encouraged on 15^th^ March 2020, schools were closed and lockdown occurred on 23^rd^ March 2020). We implement these interventions by assuming that the percentage of 0-18-year olds attending school after the 23^rd^ March 2020 was 5% (reducing all contacts between school age individuals by 95%) and that social distancing reduced all contacts by 0-50%. We take the element-wise minimum for each age group’s contact with another age group from all active interventions (distancing, schools/lockdown). A study on post-lockdown contact patterns (CoMix [11]) is used to inform contacts after lockdown (first survey 24^th^ March 2020, with an average of 73% reduction in daily contacts observed per person compared to POLYMOD).

Moving between contact matrices of multiple interventions was implemented by assuming a phased, linear decrease. After lockdown we vary a parameter (*endphase*) to capture the time taken to fully adjust (across the population, on average) to the new measures (allowed to vary from 1 to 31 days). This assumption represents the time taken for individuals to fully adapt to new measures (and household transmission), and is in line with data on the delay in the control of COVID-19 (reductions in hospital admissions and deaths after lockdown) [23]. The parameter *endphase* can be interpreted as accounting for the time taken to adjust to all interventions (and not just lockdown).

### The force of infection

The age-specific force of infection ***λ_g_***, depends on the proportion of the population who are infectious (asymptomatic ***A_g_*** and symptomatic ***I_g_*** only) and probability of transmission ***β***:

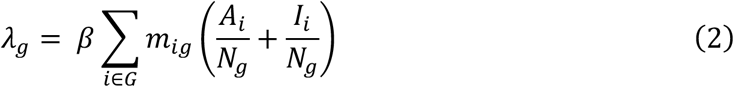

### The basic reproduction number R_0_

The basic reproduction number *R*_0_ of COVID-19 is estimated to be 2.79 *±* 1.16 [24]. We include this estimate within our model by calculating the maximum eigenvalue of the contact matrix ***M***, and allowing the transmission parameter to vary such that *R*_0_ is equal to the maximum eigenvalue of ***M*** multiplied by the infectious period *μ* and the transmission parameter *β*. This gives the value for the initial basic reproduction number *R*_0_, which changes as the contact patterns change as lockdown and other interventions are implemented.

### Parameter estimates and data sources

Model parameters are detailed in Table 1. We used available published literature to inform parameter estimates. We used the following publicly-available metrics for model fitting: regional cumulative cases in SW England (tested and confirmed case in hospital), and deaths (daily/cumulative counts) from the Public Health England COVID-19 dashboard [14], and ONS weekly provisional data on COVID-19 related deaths [25]. The case data is finalised prior to the previous five days, so we include all data until 14^th^ May 2020, based on data reported until 18^th^ May 2020. The mortality data from ONS does not explicitly state the number of COVID-19 related deaths occurring in hospital, but they do report this value nationally (83.9% of COVID-19 deaths in hospital, as of 17^th^ April 2020). We assume that this percentage applies to the SW data and re-scale the mortality to 83.9% to represent an estimate of total deaths in hospital.

**Table 1:** Parameter estimates used in the model and their sources. The distributions of unknown parameters are shown in Figure S1 for the best 100 fits.

| *Symbol* | *Description* | *Uniform prior (min and max) or point estimate* |
| --- | --- | --- |
| $1/\eta$ | Duration of the non-infectious exposure period | 5.1 days [26] |
| $\delta$ | Percentage of infections which become symptomatic | 82.1% [27]; Vary between 73.15 – 91.05% |
| $1/\mu$ | Duration of symptoms whilst not hospitalised (independent of outcome) | Vary between 2 – 14 days |
| $1/\rho$ | Duration of stay in acute bed (independent of outcome) | Vary between 2 – 14 days |
| $\gamma_g$ | Percentage of symptomatic cases which will require hospitalisation | 0-4 = 0.00%, 5-17 = 0.0408%, 18-29 = 1.04%, 30-39 = 2.04-7.00%, 40-49 = 2.53-8.68%, 50-59 = 4.86-16.7%, 60-69 = 7.01-24.0%, 70+ = 9.87-37.6% [16] |
| $1/\psi$ | Duration of stay in IC bed (independent of outcome) | 3 – 11 days [28] |
| $\epsilon$ | Percentage of those requiring hospitalisation who will require IC | Vary between 0 – 30% |
| $\omega_g$ | Percentage of those requiring IC who will die | 0-4 = 0.00%, 5-17 = 0.00%, 18-29 = 18.1%, 30-39 = 18.1%, 40-49 = 24.7%, 50-59 = 39.3%, 60-69 = 53.9%, 70+ = 65.3% [28] |
| $\kappa$ | Percentage of those requiring acute beds (but not IC) who will die | Vary between 5 – 35% |
| *school* | Percentage of 0-18-year olds attending school after 23/03/2020 | Assume 5% |
| *distancing* | Percentage reduction in contact rates due to social distancing after 15/03/2020 | Vary between 0 – 50% |
| *lockdown* | Percentage reduction in contact rates due to lockdown after 23/03/2020 | Retail/recreation: Bristol 86% Bath 90% Plymouth 85% Gloucs 84%, Somerset 82%, Devon 85%, Dorset 84% [29]<br>Transit stations: Bristol 78% Bath 71% Plymouth 65% Gloucs 69%, Somerset 67%, Devon 66%, Dorset 63% [29]<br>Vary between 63 - 90% |
| $R_0$ | Initial reproductive number of COVID-19 | 1.63 – 3.95 [24] |

### Model calibration

Using the available data (Table 1), we define ranges for all parameters in our model and sample all parameters simultaneously between these minimum and maximum values assuming uniform distributions using Latin Hypercube Sampling (statistical method for generating random parameters from multidimensional distribution), for a total of 100,000 simulations. We used maximum likelihood estimation on total cumulative cases and cumulative deaths with a Poisson negative log likelihood calculated and summed over all observed and predicted points. For *i* observed cases *X_i_* (from data), and *i* predicted cases *Y_i_* (from simulations of the model), we select the best 100 parameter sets which maximise the log-likelihood Σ *X_i_ log*(*Y_i_*) − *Y_i_* from the total sample of 100,000 simulations. The best 100 samples were taken as part of a bias-variance trade-off (Supplementary Information, Sensitivity analysis), and the qualitative inferences would not change with other choices of sample size. For each data point (taken from cases and deaths) we calculate this log-likelihood, and weight each according to the square-root of the mean of the respective case or death data. This ensures that we are considering case and death data equally within our likelihood calculations.

### Model outputs

For each of the 100 best parameter sets we run the model until 11^th^ May 2020 and output the cumulative cases and deaths in the SW. We output the predicted proportion of the population who are infectious and who have ever been infected over time. Finally, we estimate the daily and cumulative patterns of admission to and discharge from hospital (intensive care and acute) and cumulative mortality from COVID-19.

## Results and outputs

From 100,000 simulated parameter sets, we selected the best 100 baseline model fits on the basis of agreement to the calibration data on daily confirmed COVID-19 cases and weekly mortality due to COVID-19 in SW England. The distribution of the best fitting values are shown in Figure S1a (and the priors in Figure S1b). All results are shown with median and 95% credible intervals (95%CrI).

On 11^th^ May 2020, the reported cumulative number of individuals with (lab confirmed) COVID-19 was 7,116 in SW England [14], and the most recent report on total cumulative deaths showed that 2,306 had died from COVID-19 (as of 8^th^ May 2020) [25].

### Estimating the total proportion of individuals in South West England with COVID-19

Figure 2 shows the projected numbers of exposed, recovered and infectious (asymptomatic and symptomatic infections) until lockdown measures were lessened on the 11^th^ May 2020. On this date, the model predicts that a total of 5,793 (95%CrI 2,003 – 12,051) were infectious (0.10% of the total SW England population, 95%CrI 0.04 – 0.22%). The model also predicts that a total of 189,048 (95%CrI 141,580 – 277,955) have had the virus but recovered (either asymptomatically or symptomatically), which is 3.4% (95%CrI 2.5 – 5.0%) of the SW population (not infectious and not susceptible to reinfection).

**Figure 2:**
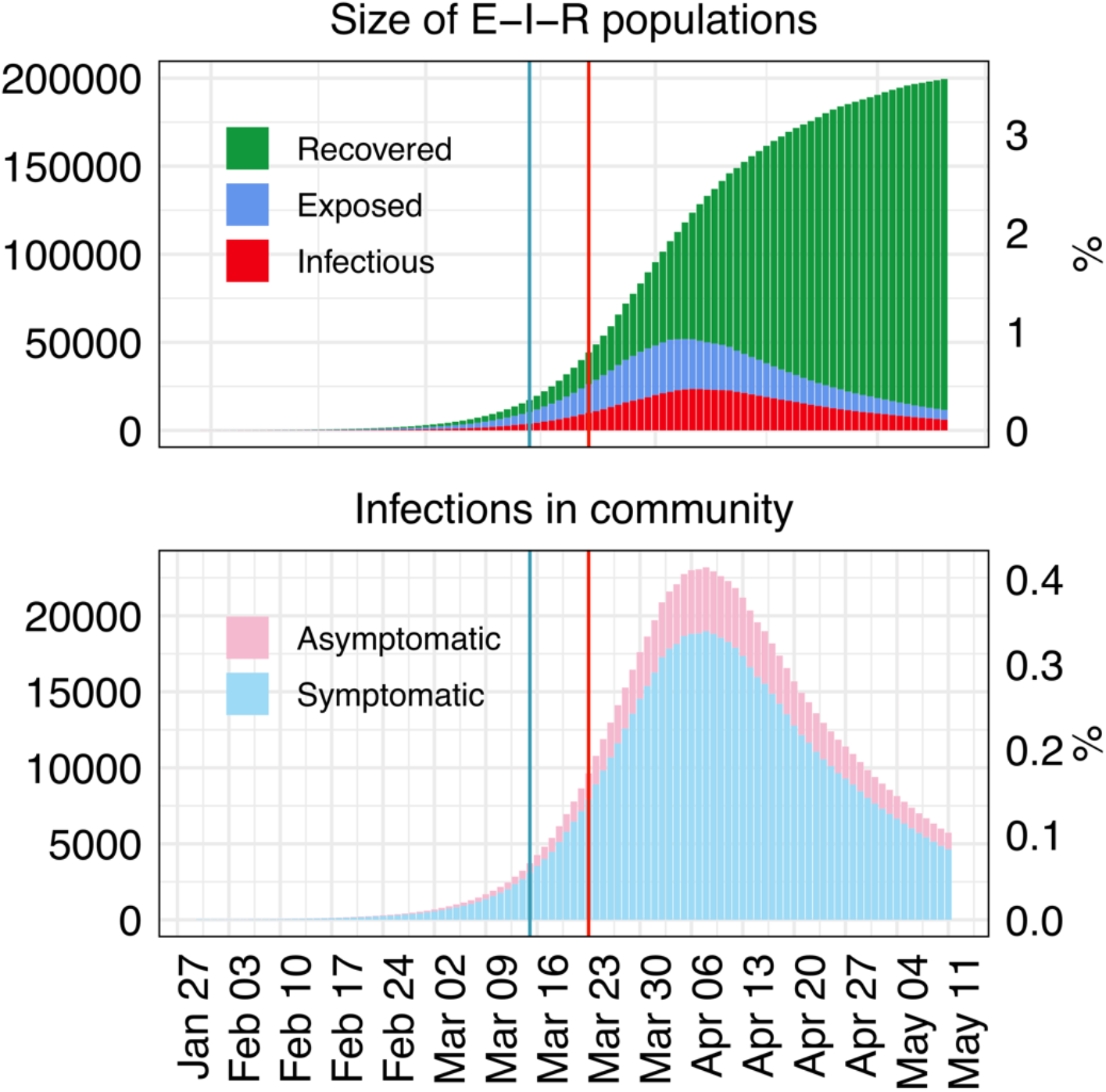
The predicted median size of the exposed (E), infectious (I) and recovered classes (R), along with the size of asymptomatic and symptomatic individuals on each day in SW England until 11^th^ May 2020. Blue and red vertical lines represent the date the government introduced social distancing and school closures/lockdown, respectively.

### Estimating the total COVID-19 hospitalised patients in acute and intensive care beds

The total number of patients in acute (non-intensive care) hospital beds across SW England was projected to be 701 (95%CrI 169 – 1,543) and the total number of patients in intensive care hospital beds was projected to be 110 (95%CrI 8 – 464) on the 11^th^ May 2020 (Figure 3). Note that these ranges are quite large due to the uncertainty in the data and as more data becomes available these predictions will change.

**Figure 3:**
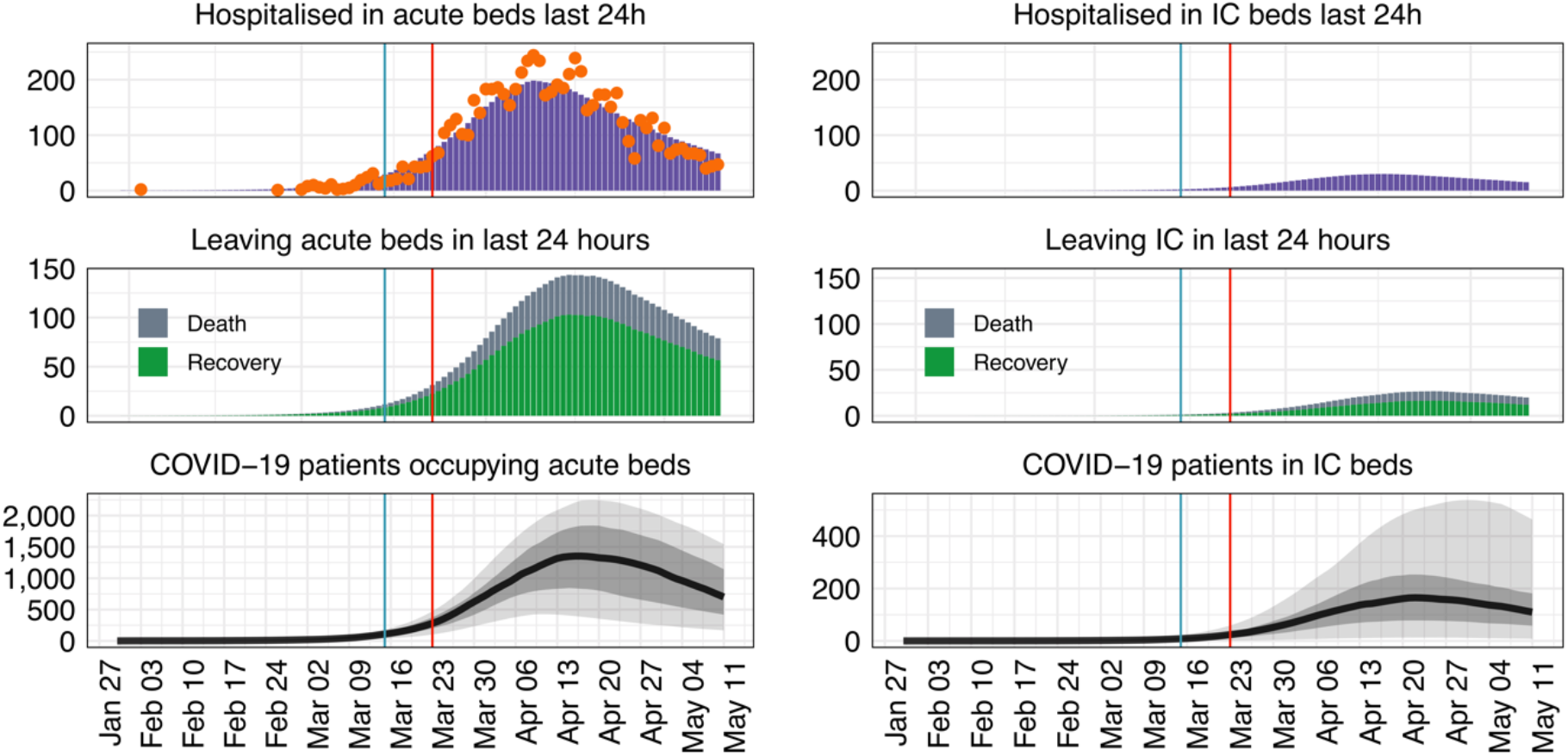
The predicted number of hospitalised patients in acute and intensive care beds in the SW until 11^th^ May 2020. The number of daily incoming patients diagnosed with COVID-19 are shown in orange (from SW daily case data [14]), 95% credible intervals are shown in light grey, 50% in dark grey and the median value of the fits is highlighted in black. The shaded region indicates the prediction of the model from the data. Blue and red vertical lines represent the date the government introduced social distancing, and school closures/lockdown, respectively.

### Estimating the reproduction number under interventions

Figure 4 shows the model prediction for the reproduction (“R”) number over time until 11^th^ May 2020, when lockdown measures were relaxed. All interventions (social distancing, school closures/lockdown) had a significant impact on the reproductive number for COVID-19 in the South West of England. We predict that prior to any interventions R was 2.6 (95%CrI 2.0 – 3.2), and the introduction of social distancing reduced this number to 2.3 (95%CrI 1.8 – 2.9). At the minimum, R was 0.6 (95%CrI 0.5 – 0.7) after all prior interventions were enacted and adhered to (social distancing, school closures and lockdown).

**Figure 4:**
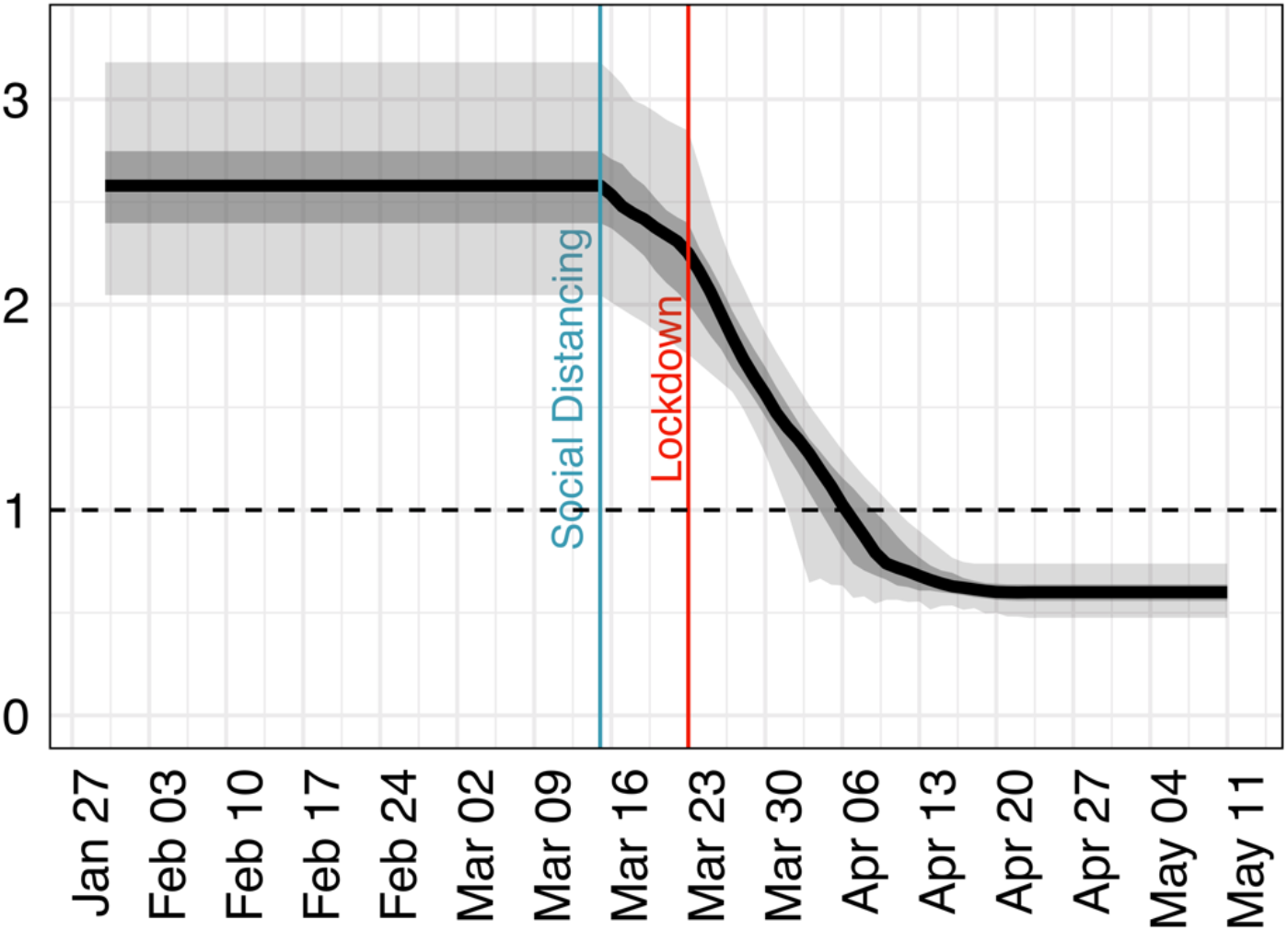
The effect of interventions on estimates of R (y-axis) over time until the 11^th^ May 2020.

## Discussion

We have developed a deterministic ordinary differential equation model of the epidemic trajectory of COVID-19 focussing on acute and IC hospital bed capacity planning to support local NHS authorities, calibrating to SW-specific data. The model is age-structured and includes time-specific implementation of current interventions (following advice and enforcement of social distancing, school closures and lockdown) to predict the potential range of COVID-19 epidemic trajectories.

Using the publicly-available data on cases and deaths, combined with the early estimates of parameters from early epidemics in other settings, we predict that on the 11^th^ May 2020 a total of 5,793 (95%CrI 2,003 – 12,051) were infectious, which equates to 0.10% (95%CrI 0.04 – 0.22%) of the total SW population. In addition, we find that the model predicts a total of 189,048 (95%CrI 141,580 – 277,955) have had the virus but recovered, which is 3.4% (95%CrI 2.5 – 5.0%) of the SW population.

We also estimate that the total number of patients in acute hospital beds in SW England on 11^th^ May 2020 was 701 (95%CrI 169 – 1,543) and in IC were 110 (95%CrI 8 – 464), while the R number has decreased from 2.6 (95%CrI 2.0 – 3.2) to 0.6 (95%CrI 0.5 – 0.7) after all interventions were enacted and fully adhered to.

The fits generally agree well with both the daily case data, and the cumulative count of deaths in the SW, although the model overestimates the case data at early stages, and underestimates later on (which can see seen in Figure S2a, and a scatter plot of expected versus observed outputs in Figure S2b). This could be because we are using formal fitting methods, or from the under-reporting of cases in the early epidemic.

The primary strength of this study is that we have developed generalisable and efficient modelling code incorporating disease transmission, interventions and hospital bed demand which can be adapted for use in other regional or national scenarios, with the model available on GitHub for open review and use (*github.com/rdbooton/bricovmod*). We have worked closely with the NHS and at Clinical Commissioning Group (CCG) level to ensure the model captures key clinical features of disease management in SW hospitals and provides output data in a format relevant to support local planning. We combined local clinical expertise with detailed literature searches, to ensure reasonable parameter ranges and assumptions in the presence of high levels of parameter uncertainty.

The main challenge of this work is in balancing the urgent need locally for prediction tools which are up to date (i.e. not relying on the national trends to inform capacity planning), versus more exhaustive and robust methods for model comparison. The latter of which uses existing models and more time-consuming (but more robust) data fitting methods [30,31]. However, we believe that release of this paper and sharing of model code will facilitate multidisciplinary collaboration, rapid review and support future model comparison and uncertainty analyses [31].

As with all models of new infections there are significant parameter uncertainties. Rapidly emerging literature is exploring a wide range of biological and epidemiological factors concerning COVID-19, but due to the worldwide nature of these studies, often parameter bands are wide and may be context specific. For example, early estimates of the basic reproduction number ranged from 1.6 to 3.8 in different locations [32,33], with an early estimate of 2.4 used in UK model projections [8]. In addition, the information which informs our parameter selection is rapidly evolving as new data is made available, sometimes on a daily basis. From our initial analysis, we identified the following parameters as critical in determining the epidemic trajectory within our model – the percentage of infections which become symptomatic, the recovery time for cases which do not require hospital, the period between acute and IC occupancy, the length of stay in IC, the probability of transmission per contact and the gradual implementation of lockdown rather than immediate effect. Other parameters (such as the percentage reduction in school-age contacts from school closures) did not seem to influence the dynamic trajectory as strongly – and thus we assume point estimates for these parameters. However, for example assuming that 95% of school-age contacts are reduced as a direct result of school closures is perhaps an overestimate, and future modelling work should address these uncertainties and their impacts on the epidemic trajectory of COVID-19 (but in this case, this value was somewhat arbitrary, and the assumption was used in the absence of school-age contact survey data). More research is urgently needed to refine these parameter ranges and to validate these biological parameters experimentally.

We have also assumed that there is no nosocomial transmission of infection between hospitalised cases and healthcare workers, as we do not have good data for within-hospital transmission. However frontline healthcare staff were likely to have been infected early on in the epidemic [34], which could have implications for our predicted epidemic trajectory. Our model also assumes a closed system, which may not strictly be true due to continuing essential travel. But given that up until 11^th^ May, travel restrictions are very severe due to lockdown measures [5], any remaining inter-regional travel is likely to have minimal effects on our model outputs.

Similar to most other COVID-19 models, we use a variant on a susceptible-exposed-infectious-recovered (SEIR) structure [8–10,16,30,35,36]. We do not spatially structure the population as in other UK modelling [9, 10], but we do include age-specific mixing based on POLYMOD data [22], and the post-lockdown CoMix study [11]. We also explicitly measure the total asymptomatic infection, and the total in each of the clinically relevant hospital classes (acute or IC), which is a strength of our approach. Future models could also take into account local bed capacity within hospitals (including Nightingale centres) and accommodate the effect of demand outstripping supply leading to excess deaths, inclusive of non-hospital-based death such as is occurring within care homes. As with all modelling, we have not taken into account all possible sources of modelling misspecification. Some of these misspecifications will tend to increase the predicted epidemic period, and others will decrease it. One factor that could significantly change our predicted epidemic period is the underlying structure within the population leading to heterogeneity in the average number of contacts under lockdown e.g. key workers have high levels of contact but others are able to minimise contacts effectively, this might lead to an underestimate of ongoing transmission, but potentially an overestimate of the effect of releasing lockdown. We also know that there are important socio-economic considerations in determining people’s ability to stay at home and particularly to work from home [37].

Early UK modelling predicted the infection peak to be reached roughly 3 weeks from the initiation of severe lockdown measures, as taken by the UK government in mid-March [8]. A more recent study factoring spatial distribution of the population indicated the peak to follow in early April due to *R*_0_ reducing to below 1 in many settings in weeks following lockdown [9]. Other modelling indicated that deaths in the UK would peak in mid-late April; furthermore, that the UK would not have enough acute and IC beds to meet demand [38]. While modelling from the European Centre for Disease Prevention and Control estimated peak cases to occur in most Euopean countries in mid-April [20], these estimations were largely at a national level. Due to the expected lag of other regions behind London, these estimated peaks are likely to be shifted further into the future for the separate regions of the UK, and as shown by our model occurred in early- to mid-April. This is also likely to be true for future peaks which may result from relaxing lockdown.

Outside of the UK, similar modelling from France [35] (which went into lockdown at a similar time the UK on 17^th^ March), predicted the peak in daily IC admissions at the end of March. Interestingly however, when dissected by region, the peak in IC bed demand varied by roughly 2 weeks. Swiss modelling similarly predicted a peak in hospitalisation and numbers of patients needing IC beds in early April, after lockdown implementation commenced on 17^th^ March [36]. US modelling [39] disaggregated by State, also highlights the peak of excess bed demand varies geographically, with this peak ranging between the 2^nd^ week of April, through to May dependent on the State under consideration. The modelling based in France also cautioned that due to only 5.7% of the population having been infected by 11^th^ May when the restrictions would be eased, the population would be vulnerable to a second epidemic peak thereafter [35].

The ONS in England estimated that an average of 0.25% of the population had COVID-19 between the 4^th^ and 17^th^ May 2020 (95% confidence interval: 0.16 – 0.38%) [40], which is greater than the 0.10% (95%CrI 0.04 – 0.22%) we found with our model (on 11^th^ May 2020), but with some overlap. In addition, the ONS estimated that 6.78% (95%CrI 5.21 – 8.64%) tested positive for antibodies to COVID-19 up to 24^th^ May 2020 in England [41], and Public Health England estimated that approximately 4% (2 – 6%) tested positive for antibodies to COVID-19 between the 20th and 26th April 2020 in the SW [42]. Comparing to our model, 3.4% (95%CrI 2.5 – 5.0%) had recovered on the 11^th^ May 2020 (2 weeks later), demonstrating that our model estimates may be within sensible bounds, and further highlighting the need for more regional estimates of crucial epidemiological parameters and seroprevalence. We have assumed that individuals are not susceptible to reinfection within the model timeframe, however in future work it will be important to explore this assumption. It is not known what the long term pattern of immunity to COVID-19 will be [43], and this will be key to understanding the future epidemiology in the absence of a vaccine or effective treatment options.

With this in mind, our findings demonstrate that there are still significant data gaps – and in the absence of such data, mathematical models can provide a valuable asset for local and regional healthcare services. This regional model will be used further in the SW as the pandemic evolves and could be used within other healthcare systems in other geographies to support localised predictions. Controlling intervention measures at a more local level could be made possible through monitoring and assessment at the regional level through a combination of clinical expertise and local policy, guided by localised predictive forecasting as presented in this study.

## Data Availability

The model was developed in R and all code and links to source data are freely available (github.com/rdbooton/bricovmod).
We used the following publicly-available metrics for model fitting: regional cumulative cases in SW England (tested and confirmed case in hospital), and deaths (daily/cumulative counts) from the Public Health England COVID-19 dashboard, and Office for National Statistics weekly provisional data on COVID-19 related deaths.

https://www.ons.gov.uk/peoplepopulationandcommunity/birthsdeathsandmarriages/deaths/datasets/weeklyprovisionalfiguresondeathsregisteredinenglandandwales

https://coronavirus.data.gov.uk/#

## Author contributions

Conception and design of the study: LD, EBP. Acquisition of data: RDB, EBP, RW, KMET. Mathematical modelling: RDB, LM, LD, EBP, RW, KMET. Coding and simulations: RDB. Analysis and interpretation of results: RDB, LM, LV, KJL, CH, PB, IH, RL, FH, DL, LD, AP, EBP, RW, KMET. Writing and drafting of the manuscript: RDB, LM, LV, KJL, CH, PB, IH, RL, FH, DL, LD, AP, EBP, RW, KMET. Approval of the submitted manuscript: RDB, LM, LV, KJL, CH, PB, IH, RL, FH, DL, LD, AP, EBP, RW, KMET.

## Data sharing statement

All model code is open source and available for download on GitHub https://github.com/rdbooton/bricovmod. All data is freely available via the GOV.UK COVID-19 dashboard [14] and ONS [21, 25].

## Supplementary information

**Table S1:**
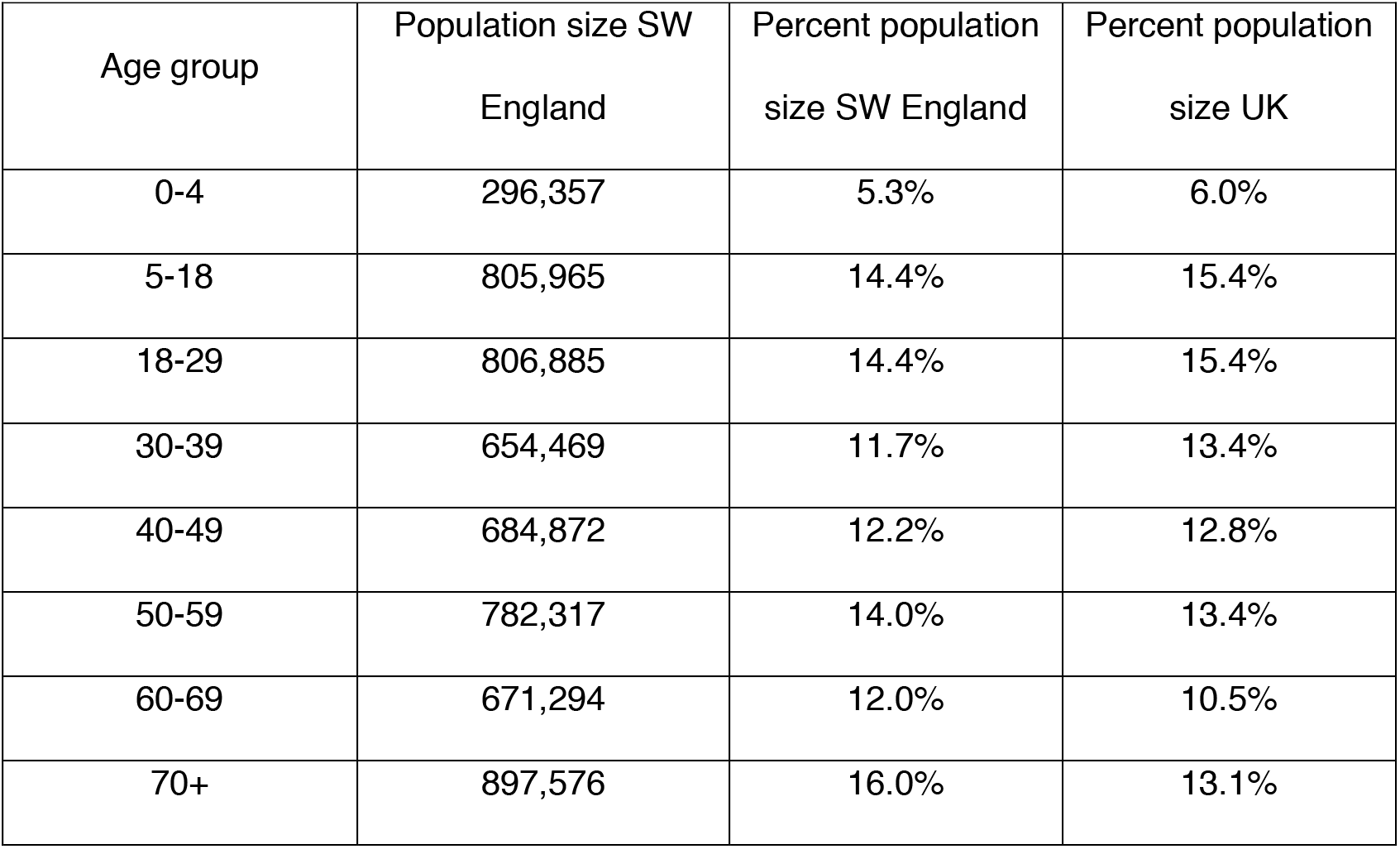
Demography of SW England compared to the UK.

| Age group | Population size SW<br>England | Percent population<br>size SW England | Percent population<br>size UK |
| --- | --- | --- | --- |
| 0-4 | 296,357 | 5.3% | 6.0% |
| 5-18 | 805,965 | 14.4% | 15.4% |
| 18-29 | 806,885 | 14.4% | 15.4% |
| 30-39 | 654,469 | 11.7% | 13.4% |
| 40-49 | 684,872 | 12.2% | 12.8% |
| 50-59 | 782,317 | 14.0% | 13.4% |
| 60-69 | 671,294 | 12.0% | 10.5% |
| 70+ | 897,576 | 16.0% | 13.1% |

### Key assumptions used in the model

i. Closed, static population size with no immigration or emigration due to the model being run over a short period of time, and current travel restrictions which should prevent significant movement of individuals in and out of the South West.
ii. No nosocomial transmission.
iii. Recovered individuals are not susceptible to reinfection within the timeframe of the model horizon.
iv. Trajectory of model outputs is assuming that there is no easing of lockdown situation over the timeframe of the model horizon.
v. There is no restriction on number of hospital (IC and acute) beds available.
vi. Asymptomatic and symptomatic are equally infectious and recover at the same rate.
vii. 95% reduction in 0- 18-year old contacts from school closures
viii. Range of 0 - 50% reduction in contacts from social distancing and range of 63 - 90% reduction in contact rates due to lockdown
ix. The reduction of any given contact rate is taken to be the minimum from lockdown, school closures and social distancing.
x. R_0 is sampled between 2.79 +/- 1.16 and the infectious period and transmission probability are chosen to achieve this
xi. Proportion symptomatic which require hospitalisation depends on age, with increased risk for older age groups
xii. Proportion in IC who will die depends on age, with increased risk for older age groups
xiii. The percentage of infections which become symptomatic is 73.15 - 91.05%
xiv. The percentage of those requiring hospital who will require IC 0 - 30%
xv. The percentage of those requiring acute beds (but not IC) who will die 5 - 35%

**Figure S1a:**
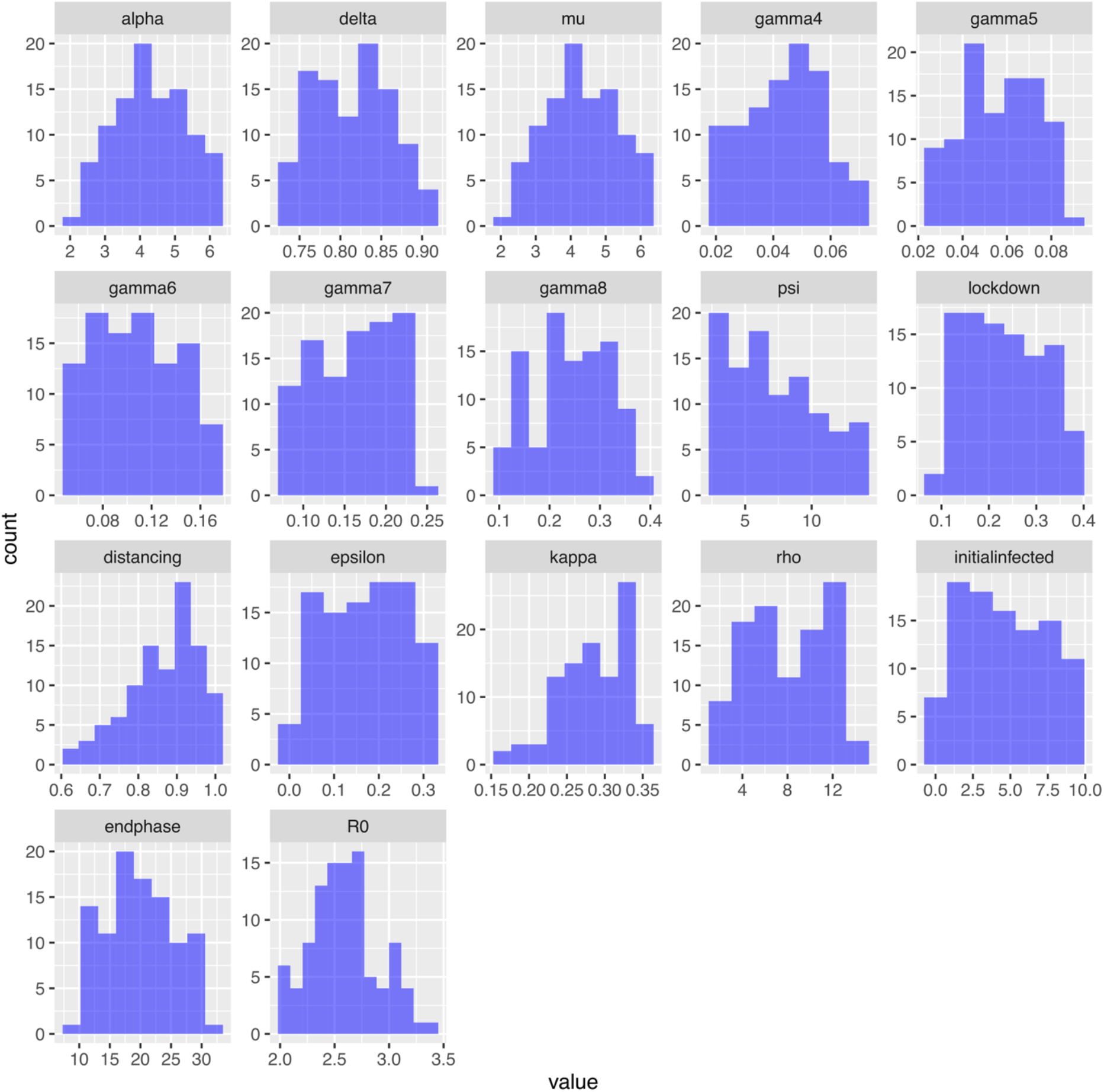
The distribution of the best 100 parameters selected for in the Latin Hypercube Sampling and likelihood fitting from 100,000 simulations.

**Figure S1b:**
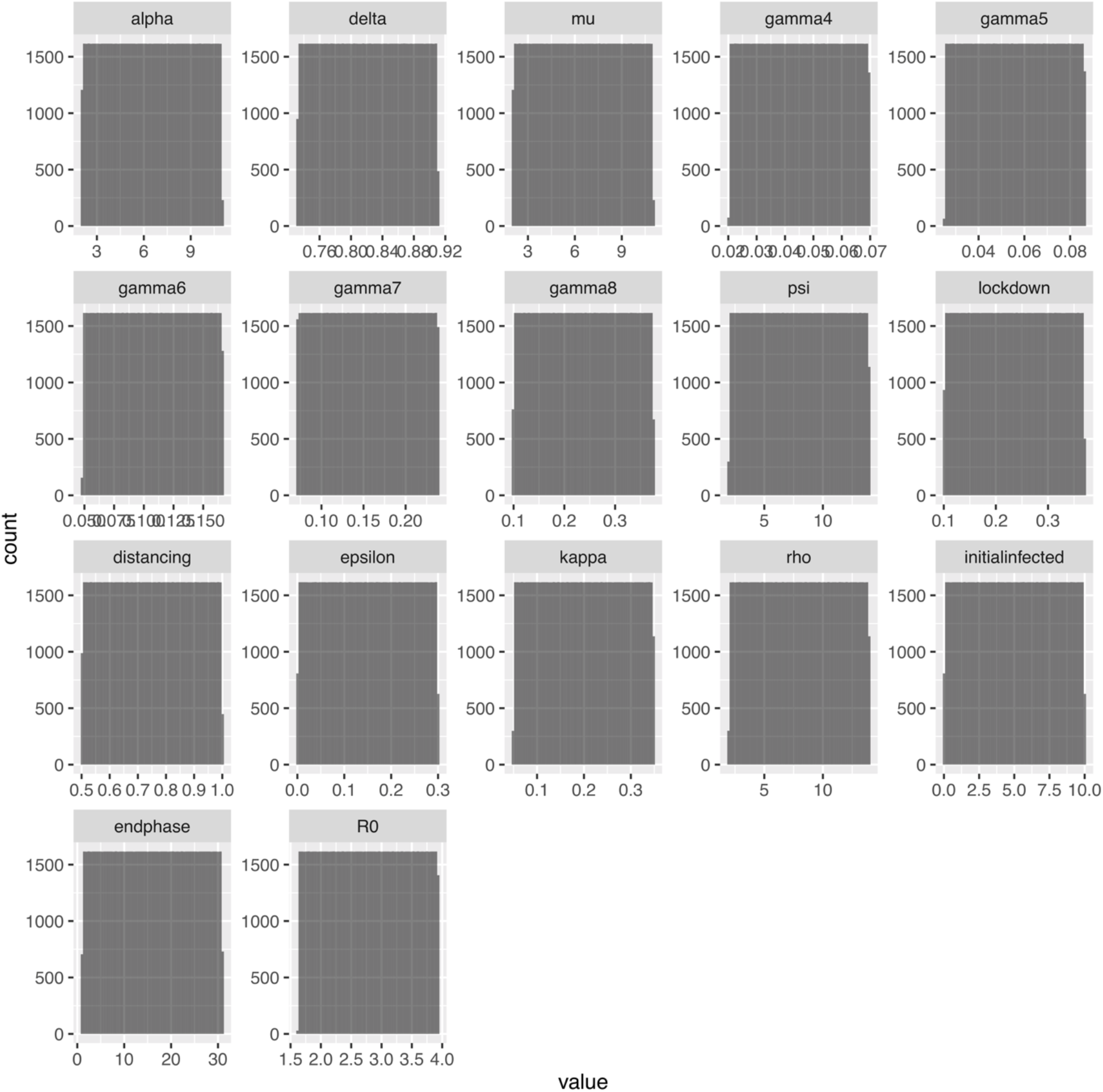
The distribution of the uniform priors in the Latin Hypercube Sampling from 100,000 samples.

**Figure S2a:**
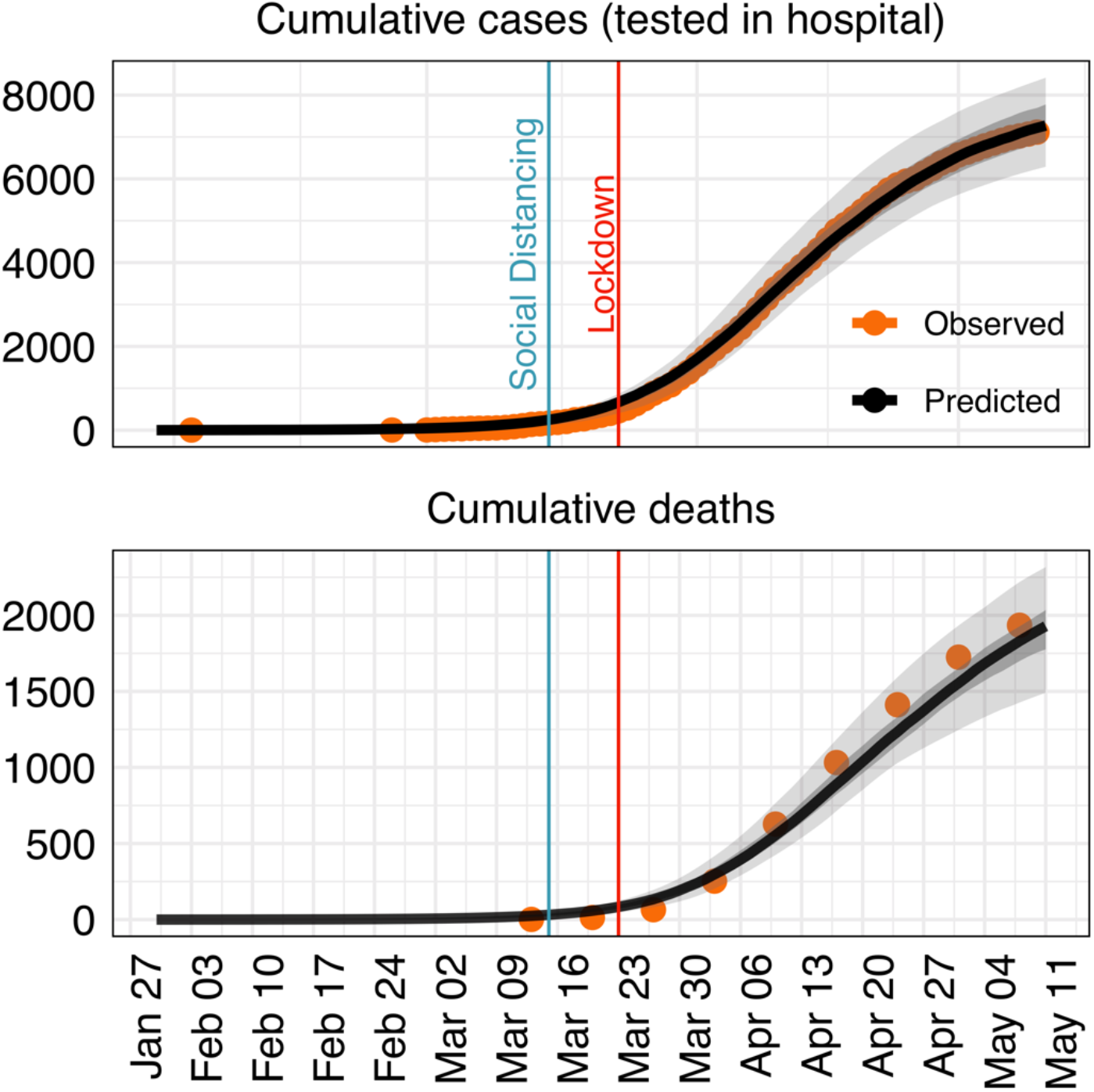
Fitting performance of the model. The cumulative case numbers and hospital deaths for COVID-19 in SW England until lockdown measures were gradually lifted (11^th^ May 2020), based on case data and death data (orange). 95% credible intervals of our model projections are shown in light grey, 50% in dark grey and the median value of the model is highlighted in black. The shaded region indicates the prediction of the model from the data. We did not consider the effects of lockdown being lifted, and our transmission rates are fitted to both before lockdown and after lockdown. Blue and red vertical lines represent the dates when social distancing and school closures/lockdown were introduced nationally, respectively.

**Figure S2b:**
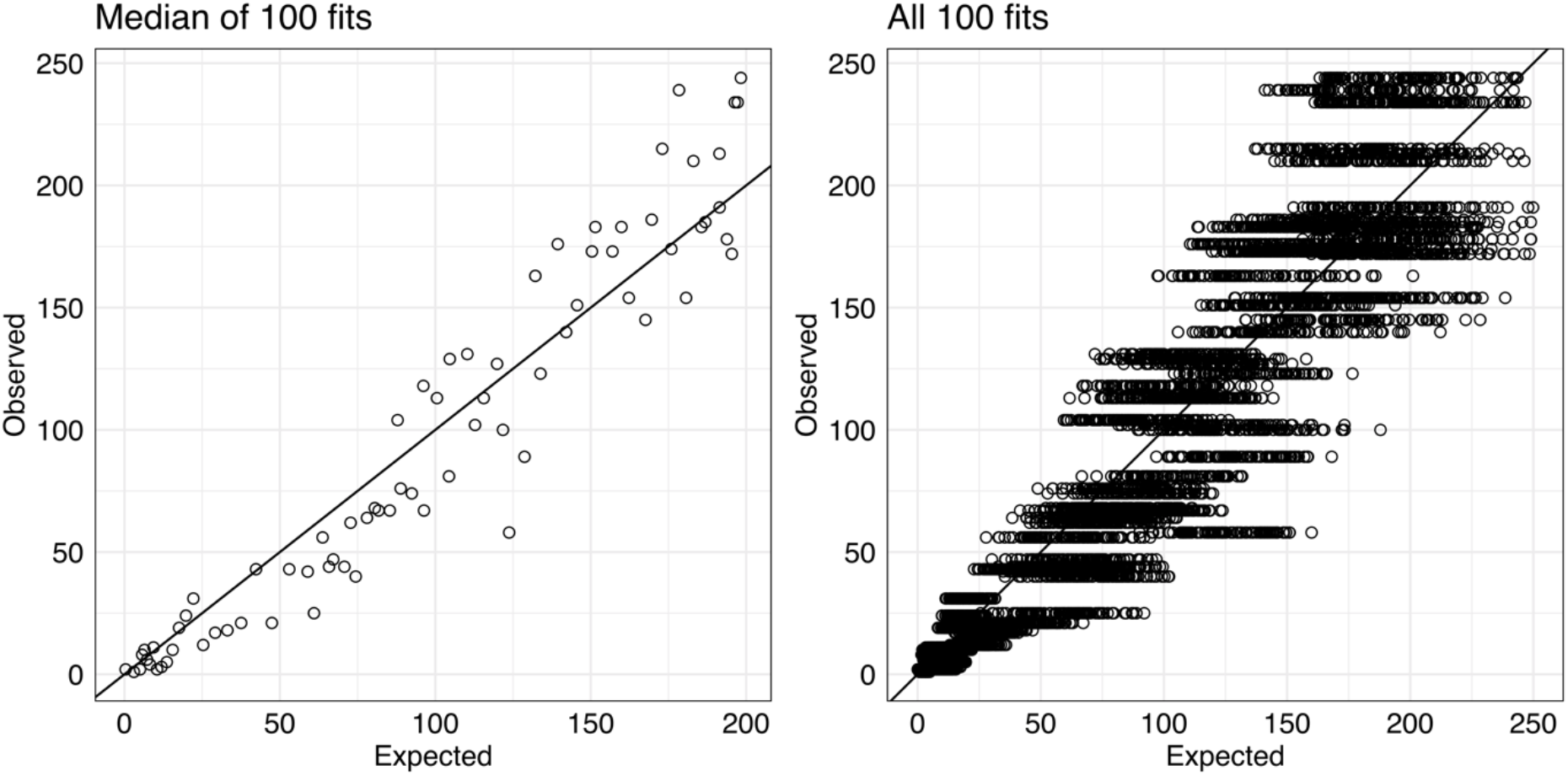
The expected (model) versus observed (data) for the median of 100 fits, and all 100 fits for counts of daily case numbers. The model tends to overestimate the case data at earlier stages of the epidemic and underestimate at later stages of the epidemic.

## Sensitivity analysis

The choice of the final sample size *m*=100 taken from 100k samples is based on a bias-variance trade-off with lower m giving a better model fit but higher *m* allowing more predictions to be included. The choice of *m* is not of practical significance for the median and 95% credible intervals of the main outcome measures of the model – bed modelling - as shown in the below table.

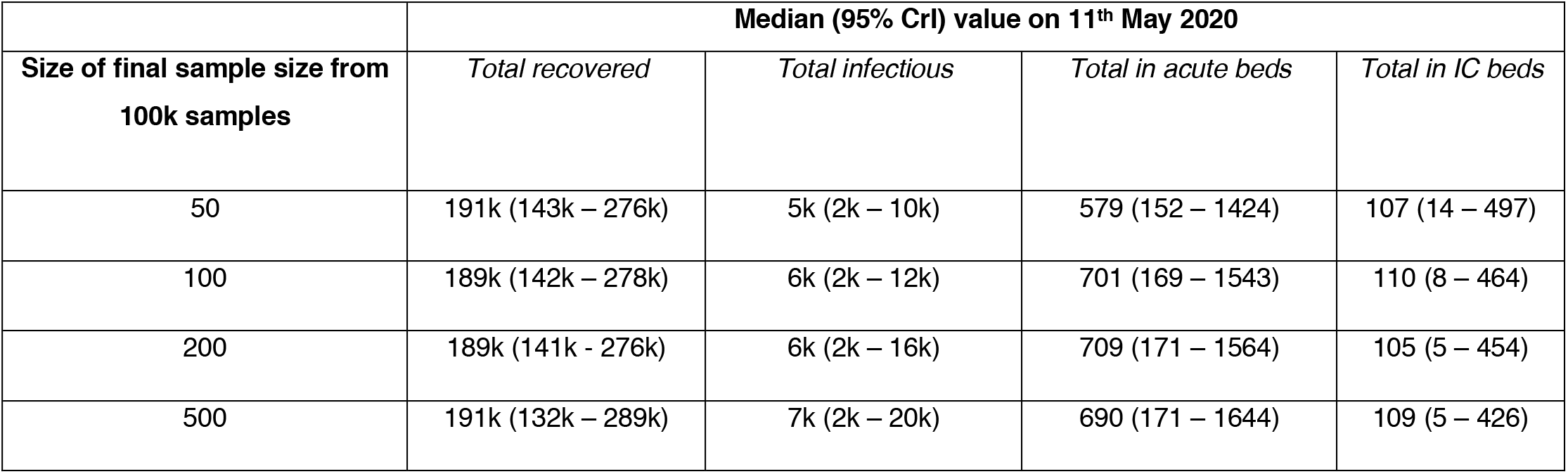

| Size of final sample size from 100k samples | Median (95% CrI) value on 11<sup>th</sup> May 2020 |  |  |  |
| --- | --- | --- | --- | --- |
|  | *Total recovered* | *Total infectious* | *Total in acute beds* | *Total in IC beds* |
| 50 | 191k (143k – 276k) | 5k (2k – 10k) | 579 (152 – 1424) | 107 (14 – 497) |
| 100 | 189k (142k – 278k) | 6k (2k – 12k) | 701 (169 – 1543) | 110 (8 – 464) |
| 200 | 189k (141k – 276k) | 6k (2k – 16k) | 709 (171 – 1564) | 105 (5 – 454) |
| 500 | 191k (132k – 289k) | 7k (2k – 20k) | 690 (171 – 1644) | 109 (5 – 426) |

Increasing *m* does slightly increase the estimate of the infectious population (*total infectious*). This is explained by including models with slightly higher *R* value, i.e. *R* is biased upwards as we move further away from the best answer. The bias is small compared to the modelling uncertainty.

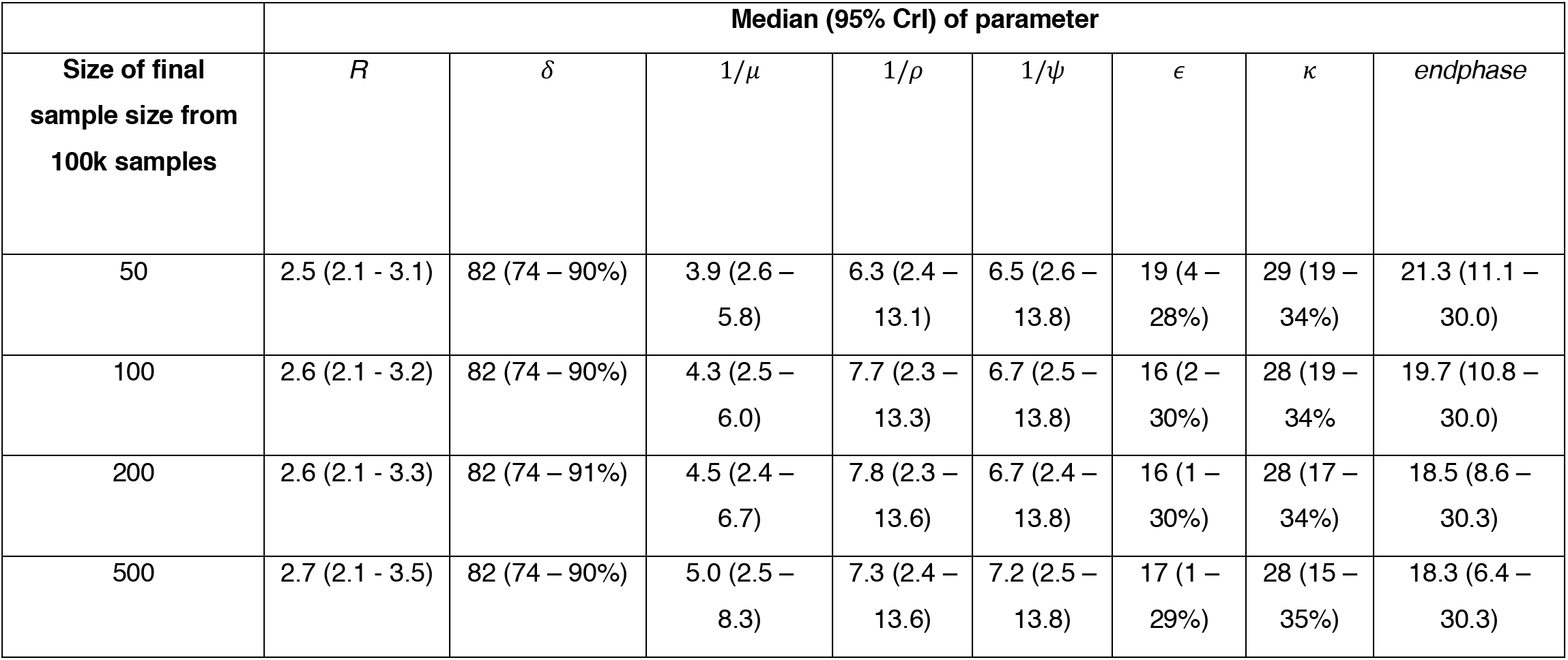

The inverse of *μ* and *ψ* becomes bigger (as it is related to the selection of R), as the sample size becomes larger. This then reduces the parameters *endphase, κ*, and ∊ in order to account for this bias in R.

We choose to report values for *m*=100 samples as part of the bias-variance trade-off. The bias is small for this choice, with R close to unbiased and the confidence intervals capture all of the significant variation components. The qualitative inferences would not change with any of the above choices of *m* and uncertainty has been well captured.

## REFERENCES

1. World Health Organization. WHO Director-General’s opening remarks at the mission briefing on COVID-19. 2020 [cited 27 Apr 2020]. Available: https://www.who.int/dg/speeches/detail/who-director-general-s-opening-remarks-at-the-media-briefing-on-covid-19---11-march-2020

2. Johns Hopkins University. COVID-19 Data Center. COVID-19 Case Tracker. [cited 27 Apr 2020]. Available: https://coronavirus.jhu.edu/

3. Payne C. State intervention in China. Nat Hum Behav. 2020;1.

4. Public Health Agency of Sweden. Public Health Authority regulations and general advice on everyone’s responsibility to prevent the infection of COVID-19. 2020 [cited 7 Apr 2020]. Available: https://www.folkhalsomyndigheten.se/publicerat-material/publikationsarkiv/h/hslf-fs-202012/

5. Public Health England. Guidance on social distancing for everyone in the UK. In: Guidance on social distancing for everyone in the UK [Internet]. 2020 [cited 27 Apr 2020]. Available: https://www.gov.uk/government/publications/covid-19-guidance-on-social-distancing-and-for-vulnerable-people/guidance-on-social-distancing-for-everyone-in-the-uk-and-protecting-older-people-and-vulnerable-adults

6. Prem K, Liu Y, Russell TW, Kucharski AJ, Eggo RM, Davies N, et al. The effect of control strategies to reduce social mixing on outcomes of the COVID-19 epidemic in Wuhan, China: a modelling study. Lancet Public Heal. 2020.

7. Flaxman S, Mishra S, Gandy A, Unwin JT, Coupland H, Mellan TA, et al. Estimating the number of infections and the impact of non-pharmaceutical interventions on COVID-19 in 11 European countries. Imp Coll London COVID-19 Report No 13. 2020.

8. Ferguson NM, Laydon D, Gemma Nedjati-Gilani, Natsuko Imai, Kylie Ainslie MB, Sangeeta Bhatia, Adhiratha Boonyasiri, Zulma Cucunubá, Gina Cuomo-Dannenburg, Amy Dighe I, Dorigatti, Han Fu, Katy Gaythorpe, Will Green, Arran Hamlet, Wes Hinsley, Lucy C Okell S van, Elsland, Hayley Thompson, Robert Verity, Erik Volz, Haowei Wang, Yuanrong Wang PGW, et al. Impact of non-pharmaceutical interventions (NPIs) to reduce COVID19 mortality and healthcare demand. Imp Coll London COVID-19 Report No 9. 2020.

9. Challen R, Tsaneva-Atanasova K, Pitt M. Estimates of regional infectivity of COVID-19 in the United Kingdom following imposition of social distancing measures. medRxiv. 2020.

10. Danon L, Brooks-Pollock E, Bailey M, Keeling MJ. A spatial model of CoVID-19 transmission in England and Wales: early spread and peak timing. medRxiv. 2020.

11. Jarvis CI, Zandvoort K Van, Gimma A, Prem K, Group CC-19 working, Klepac P, et al. Quantifying the impact of physical distance measures on the transmission of COVID-19 in the UK. medRxiv. 2020.

12. The Novel Coronavirus Pneumonia Emergency Response Epidemiology Team. The epidemiological characteristics of an outbreak of 2019 novel coronavirus diseases (COVID-19) — China, 2020. Chin J Epidemiol. 2020;41.

13. Onder G, Rezza G, Brusaferro S. Case-fatality rate and characteristics of patients dying in relation to COVID-19 in Italy. JAMA - Journal of the American Medical Association. 2020.

14. Public Health England. Coronavirus (COVID-19) in the UK: data dashboard. 2020 [cited 28 Apr 2020]. Available: https://coronavirus.data.gov.uk/#

15. Yang X, Yu Y, Xu J, Shu H, Xia J, Liu H, et al. Clinical course and outcomes of critically ill patients with SARS-CoV-2 pneumonia in Wuhan, China: a single-centered, retrospective, observational study. Lancet Respir Med. 2020.

16. Verity R, Okell LC, Dorigatti I, Winskill P, Whittaker C, Imai N, et al. Estimates of the severity of coronavirus disease 2019: a model-based analysis. Lancet Infect Dis. 2020.

17. McCurdy C. Ageing, fast and slow: when place and demography collide. Resolut Found. 2019. Available: https://www.resolutionfoundationorg/app/uploads/2019/10/Ageing-fast-and-slowpdf

18. Yang J, Zheng Y, Gou X, Pu K, Chen Z, Guo Q, et al. Prevalence of comorbidities in the novel Wuhan coronavirus (COVID-19) infection: a systematic review and meta-analysis. Int J Infect Dis. 2020.

19. Zhou F, Yu T, Du R, Fan G, Liu Y, Liu Z, et al. Clinical course and risk factors for mortality of adult inpatients with COVID-19 in Wuhan, China: a retrospective cohort study. Lancet. 2020.

20. Stockholm: ECDC (European Centre for Disease Prevention and Control). Coronavirus disease 2019 (COVID-19) pandemic: increased transmission in the EU/EEA and the UK – seventh update, 25 March 2020. 2020. Available: https://www.ecdc.europa.eu/sites/default/files/documents/RRA-seventh-update-Outbreak-of-coronavirus-disease-COVID-19.pdf

21. Office for National Statistics. Estimates of the population for the UK, England and Wales, Scotland and Northern Ireland. 2020. Available: https://www.ons.gov.uk/peoplepopulationandcommunity/populationandmigration/populationestimates/datasets/populationestimatesforukenglandandwalesscotlandandnorthernireland

22. Mossong J, Hens N, Jit M, Beutels P, Auranen K, Mikolajczyk R, et al. Social contacts and mixing patterns relevant to the spread of infectious diseases. PLoS Med. 2008. doi:10.1371/journal.pmed.0050074

23. Stedman M, Davies M, Anderson SG, Lunt M, Verma A, Heald A. A phased approach to unlocking during the COVID-19 pandemic; Lessons from trend analysis. medRxiv. 2020. doi:10.1101/2020.04.20.20072264

24. Liu Y, Gayle AA, Wilder-Smith A, Rocklöv J. The reproductive number of COVID-19 is higher compared to SARS coronavirus. J Travel Med. 2020;27. doi: 10.1093/jtm/taaa021

25. Office for National Statistics. Deaths registered in England and Wales, provisional 2020: up to week ending 17 April 2020. 2020. Available: https://www.ons.gov.uk/peoplepopulationandcommunity/birthsdeathsandmarriages/deaths/datasets/weeklyprovisionalfiguresondeathsregisteredinenglandandwales

26. Lauer SA, Grantz KH, Bi Q, Jones FK, Zheng Q, Meredith HR, et al. The incubation period of coronavirus disease 2019 (COVID-19) from publicly reported confirmed cases: estimation and application. Ann Intern Med. 2020.

27. Mizumoto K, Kagaya K, Zarebski A, Chowell G. Estimating the asymptomatic proportion of coronavirus disease 2019 (COVID-19) cases on board the Diamond Princess cruise ship, Yokohama, Japan, 2020. Eurosurveillance. 2020.

28. ICNARC COVID-19 study case mix programme database. ICNARC report on COVID-19 in critical care 15 May 2020. 2020. Available: https://www.icnarc.org/Our-Audit/Audits/Cmp/Reports

29. Google. United Kingdom COVID-19 Community Mobility Report: Mobility changes April 5 2020. 2020. Available: https://www.gstatic.com/covid19/mobility/2020-04-05_GB_Mobility_Report_en.pdf

30. Wynants L, Van Calster B, Bonten MMJ, Collins GS, Debray TPA, De Vos M, et al. Prediction models for diagnosis and prognosis of covid-19 infection: systematic review and critical appraisal. BMJ. 2020.

31. Latif S, Usman M, Manzoor S, Iqbal W, Qadir J, Tyson G, Castro I, Razi A, Boulos MN CJ. Leveraging Data Science To Combat COVID-19: A Comprehensive Review. 2020.

32. Kucharski AJ, Russell TW, Diamond C, Liu Y, Edmunds J, Funk S, et al. Early dynamics of transmission and control of COVID-19: a mathematical modelling study. Lancet Infect Dis. 2020.

33. Read JM, Bridgen JRRE, Cummings DATA, Ho A, Jewell CP. Novel coronavirus 2019-nCoV: early estimation of epidemiological parameters and epidemic predictions. medRxiv. 2020.

34. The Centre for Evidence-Based Medicine Website. How many healthcare workers are infected? 2020 [cited 23 Apr 2020]. Available: https://www.cebmnet/covid-19/covid-19-how-many-healthcare-workers-are-infected/

35. Salje H, Kiem C, Lefrancq N, Courtejoie N, Bosetti P. Estimating the burden of SARS-CoV-2 in France. HAL ID pasteur-02548181. 2020.

36. Althaus C. Real-time modeling and projections of the COVID-19 epidemic in Switzerland. Inst Soc Prev Med Univ Bern, Switz. 2020. Available: https://ispmberngithubio/covid-19/swiss-epidemic-model/

37. Dingel J, Neiman B. How Many Jobs Can be Done at Home? NBER Work Pap. 2020.

38. Institute for Health Metrics and Evaluation. New COVID-19 forecasts for Europe: Italy & Spain have passed the peak of their epidemics; UK, early in its epidemic, faces a fast-mounting death toll. 2020. Available: http://www.healthdata.org/news-release/new-covid-19-forecasts-europe-italy-spain-have-passed-peak-their-epidemics-uk-early-its

39. IHME COVID-19 health service utilization forecasting team. Forecasting COVID-19 impact on hospital bed-days, ICU-days, ventilator-days and deaths by US state in the next 4 months. medxriv. 2020.

40. ONS. Coronavirus (COVID-19) Infection Survey pilot: England, 21 May 2020. 2020.

41. ONS. Coronavirus (COVID-19) Infection Survey pilot: 28 May 2020. 2020.

42. Public Health England. Weekly Coronavirus Disease 2019 (COVID-19) Surveillance Report, Week 22. 2020. Available: https://assets.publishing.service.gov.uk/government/uploads/system/uploads/attachment_data/file/888254/COVID19_Epidemiological_Summary_w22_Final.pdf

43. Altmann DM, Douek DC, Boyton RJ. What policy makers need to know about COVID-19 protective immunity. The Lancet Comments. 2020.

